# Early indirect impact of COVID-19 pandemic on utilization and outcomes of reproductive, maternal, newborn, child and adolescent health services in Kenya

**DOI:** 10.1101/2020.09.09.20191247

**Authors:** Duncan Shikuku, Irene Nyaoke, Sylvia Gichuru, Onesmus Maina, Martin Eyinda, Pamela Godia, Lucy Nyaga, Charles Ameh

## Abstract

**Background:** The COVID-19 global pandemic is expected to result in 8.3–38.6% additional maternal deaths in many low-income countries. The objective of this paper was to determine the initial impact of COVID-19 pandemic on reproductive, maternal, newborn, child and adolescent health (RMNCAH) services in Kenya.

**Methods:** Data for the first four months (March-June) of the pandemic and the equivalent period in 2019 were extracted from Kenya Health Information System. Two-sample test of proportions for hospital attendance for select RMNCAH services between the two periods were computed.

**Results:** There were no differences in monthly mean (±SD) attendance between March-June 2019 vs 2020 for antenatal care (400,191.2±12,700.0 vs 384,697.3±20,838.6), hospital births (98,713.0±4,117.0 vs 99,634.5±3,215.5), family planning attendance (431,930.5±19,059.9 vs 448,168.3±31,559.8), post-abortion care (3,206.5±111.7 vs 448,168.3±31,559.8) and pentavalent 1 immunisation (114,701.0±3,701.1 vs 110,915.8±7,209.4), p>0.05. However, there were increasing trends for adolescent pregnancy rate, significant increases in FP utilization among young people (25.7% to 27.0%), injectable (short-term) FP method uptake (58.2% to 62.3%), caesarean section rate (14.6% to 15.8%), adolescent maternal deaths (6.2% to 10.9%) and fresh stillbirths (0.9% to 1.0%) with a reduction in implants (long-term) uptake (16.5% to 13.0%) (p< 0.05). No significant change in maternal mortality ratio between the two periods (96.6 vs 105.8/100,000 live births, p = 0.1023) although the trend was increasing.

**Conclusion:** COVID-19 may have contributed to increased adolescent pregnancy, adolescent maternal death and stillbirth rates in Kenya. If this trend persists, recent gains achieved in maternal and perinatal health in Kenya will be lost. With uncertainty around the duration of the pandemic, strategies to mitigate against catastrophic indirect maternal health outcomes are urgently needed.

## Introduction

On March 11, 2020, the World Health Organization (WHO) declared COVID-19 disease caused by the novel coronavirus (SARS-CoV-2) a global pandemic [1]. Globally, 10,185,374 cases of COVID-19 were confirmed and 503,862 deaths translating to a 4.95 percent case fatality rate were reported as at June 30, 2020 [2]. As at the stated time, Africa regions had reported the lowest number of confirmed cases, case fatality and deaths – 297 290 cases, 2.02% and 6 010 deaths respectively [2]. Cases continue to increase and in Asia and Europe, a second wave is beginning to emerge [3–5]. Health systems are expected to adapt to maintain the routine essential health services in addition to managing an increasing COVID-19 case-load.

Public health emergencies have shown that the impact of an epidemic on reproductive, maternal and newborn health, domestic and gender-based violence and mental health often goes unrecognized, because the effects are often not the direct result of the infection, but instead the indirect consequences of strained health care systems, disruptions in care and redirected resources [6–8]. As noted by the World Health Organization (WHO), “People, efforts, and medical supplies all shift to respond to the emergency. This often leads to the neglect of basic and regular essential health services. People with health problems unrelated to the epidemic find it harder to get access to health care services [9].” To avert indirect morbidity and mortality when services are disrupted, WHO developed and updated guidelines to support country preparedness and response plan [10]. These included prioritization and continuation of context-relevant essential health services during the acute phase of the COVID-19 pandemic and community-based health care, including outreach and campaigns, in the context of the COVID-19 pandemic at the end of June 2020 [11, 12]. Among the high-priority categories of essential health services by WHO included - essential prevention and treatment services for communicable diseases, including immunizations; services related to reproductive health, including during pregnancy and childbirth; and provision of medications, supplies and support from health care workers for the ongoing management of chronic diseases, including mental health conditions [6].

Governments around the world have had to quickly adapt and respond to curb transmission of the virus and to provide care for the many who have been infected. National mandates to contain the pandemic, such as complete or total lockdowns, curfews, and temporal closure of non-essential services (elective survey, cancer treatment etc), the resultant economic slowdowns have adverse effects on accessing, utilising and provision of RMNCAH services especially for low and middle income countries [6]. Evidence from the 2014 outbreak of Ebola virus in west Africa showed that the indirect effects of the outbreak were more severe than the outbreak itself [13]. In Sierra Leone, maternal and newborn care service utilization decreased due to disrupted health services and fear of seeking treatment during the outbreak related to fear of contracting Ebola virus at health facilities, distrust of the health system, and rumours about the source of the disease [14]. There were reductions in antenatal care coverage by 22 percent, family planning (6 percent), facility births (8 percent), and postnatal care (13 percent) [15] contributing to an estimated 3,600 maternal deaths, neonatal deaths and stillbirths—almost similar to the deaths (about 4,000) directly caused by the Ebola virus in the country [15]. Elsewhere, there were sharp reduction in contraceptive use and family planning visits in Guinea, Liberia and Sierra Leone due to the Ebola outbreak [16, 17]. As a result, policy makers must consider not only the immediate health effects of the pandemic but also the indirect effects of the pandemic and rapidly develop strategies to mitigate these.

Statistical models in the early phases of the COVID-19 pandemic predicted a reduction in the overall utilization of reproductive, maternal and newborn health services with related adverse mortality outcomes across the globe [18, 19]. For instance, a reduction in coverage of essential maternal health interventions by 9.8 – 51.9% would result in 8.3 – 38.6% increase in maternal deaths per month [18]. Consequently, a 10% decline in use of short- and long-acting reversible contraceptives would result in 48,558,000 additional women with an unmet need for modern contraceptives and 15,401,000 additional unintended pregnancies [19]. The objective of this study was to determine the initial impact of COVID-19 pandemic on RMNCAH services in Kenya in the first four months of the pandemic by comparing RMNCAH Kenya Health Information System (KHIS) utilization data for 2019 and 2020.

## Methods

### Study Design

This was a cross – sectional design comparing RMNCAH utilization data for Kenya extracted from the national KHIS, formerly known as the district health information system version 2 (DHIS2), an open-access publicly available reporting platform managed by the Ministry of Health (MOH). This design was used as it helps to describe the status, any changes or relationships in a phenomena over a fixed point in time [20]. Four-month period from March to June 2020 was assessed for any trends in the utilization of pregnancy and antenatal care, facility/skilled childbirths, family planning, post-abortion care and immunization services. This data was compared to the equivalent period in 2019, a year before the COVID-19 pandemic, to demonstrate any significant changes in the utilization and outcomes of these services during the COVID-19 outbreak.

### Study Setting

Kenya is a low- and middle-income country in the sub-Saharan Africa located in the East Africa region. It is geographically divided into 47 counties/county governments. Overall, the national government is mandated with policy and guidelines development while the county governments are mandated to support the implementation of the national policies and guidelines.

Kenya reported its first COVID-19 case on March 13, 2020. The government advanced preparedness, readiness and preventive measures for COVID-19 pandemic as well as to address the emerging effects of COVID-19 on the health and economy. Key measures are summarized below and the periods of enforcement against number of cases and deaths are presented inFigure 1.

**Figure 1:**
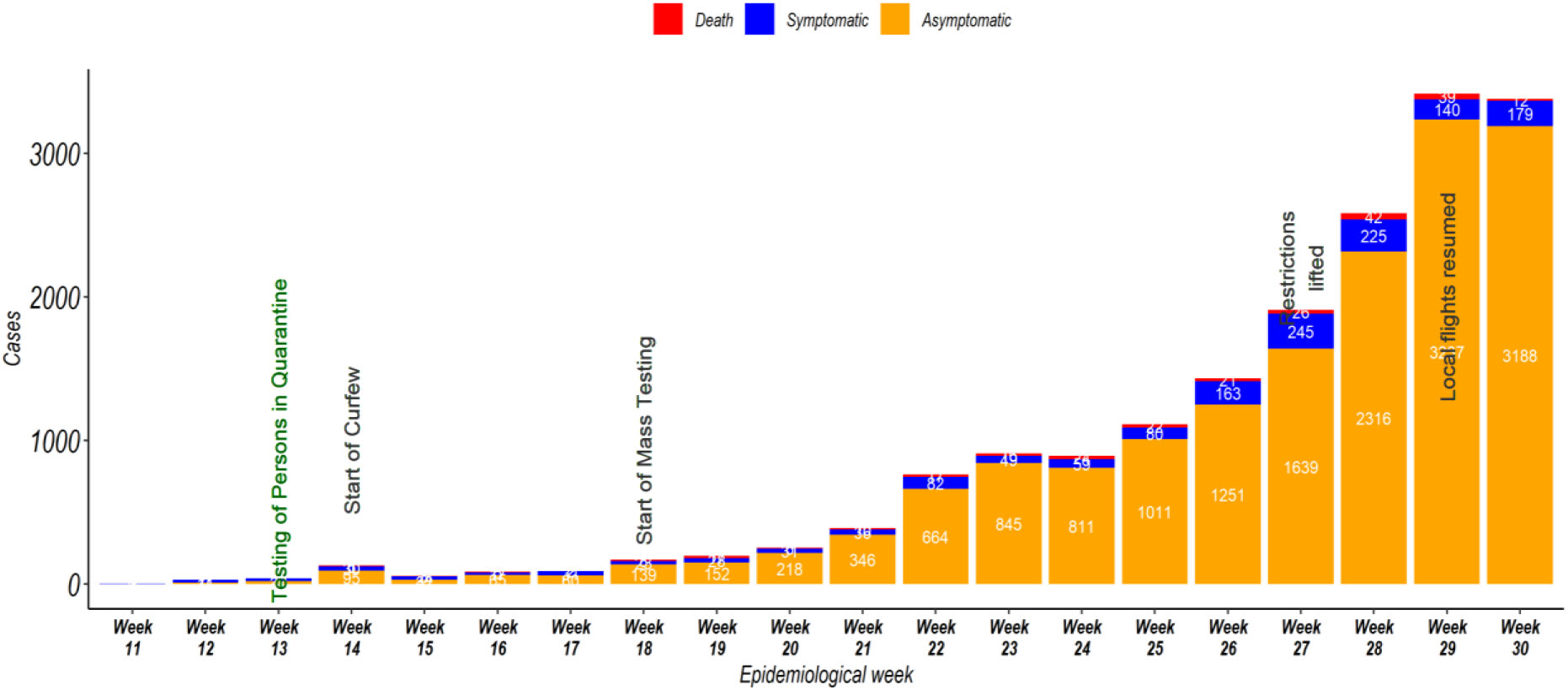
Trends of COVID-19 outbreak in Kenya and the containment measures (Adopted from the Ministry of Health (Kenya) COVID-19 Outbreak in Kenya: Daily Situation Report - 131)

March 2020: start of mandatory quarantine; testing of persons in quarantine; contact tracing, quarantine and testing of exposed cases; daily briefings by the MOH on the state and progress of COVID-19 since the first case; public awareness on COVID-19 through television, radio and print media (briefings and awareness communication ongoing at the time of writing).
April 2020: start of countrywide dawn-to-dusk curfews; partial lockdowns/restrictions of movement in and out of priority high-risk counties of Nairobi, Mombasa, Kilifi, Kwale and Mandera; restrictions to social gatherings; closure of all schools and institutions of higher learning. Other measures initiated in April were handwashing facilities in all public places; use of face masks for personal protection for all citizens; restrictions to social gatherings; enforcement of social distancing at social places including health care facilities; development and dissemination of COVID-19 guidelines - interim guidance on continuity of essential health services during the COVID-19 pandemic.
May 2020: start of mass testing; development and dissemination of guidelines - Practical Guide for Continuity of Reproductive, Maternal, Newborn and Family Planning Care and Services in the Background of COVID-19 Pandemic; establishment of the Presidential 8-point Economic Stimulus Package for COVID-19 response that included recruitment of additional 5000 frontline healthcare workers for one year, expansion of bed capacity in public hospitals and development of a welfare package to cushion frontline healthcare workers; live weekly #AskTheDG (Ask the Director General for Health) open twitter chat from early May 2020 - a platform to hear views and opinions from the public by the Director General for Health as well as to respond to medical/technical questions relating to COVID-19 in Kenya were conducted.
June 2020: launch of the Home-Based Care Isolation and Care protocol and training of health care workers and community health volunteers to educate the caregivers/households about the program aimed at building capacity at household level to take care of COVID-19 patients at home.
Early July 2020: Restrictions to movement/lockdowns lifted

As at June 30, 2020, 41 out of the 47 (87 percent) counties of Kenya had reported COVID-19 cases with exponential increases reported for the positive cases from May from the onset of mass testing [21]. In addition, a total of 6,190 confirmed cases with 144 deaths (a 2.3% case fatality rate) were reported [2].

### Data analysis

Data on RMNCAH service utilization variables of interest were extracted from the KHIS to Microsoft Excel 2016 for processing and cleaning. The variables of interest were antenatal care attendance (new and revisits; adolescent pregnancies), skilled births - total births and caesarean section rates, family planning services uptake by age and method, immunization and post-abortion care services accessed. Relevant rates/proportions were computed. Cleaned data was exported to STATA v12 (StataCorp. 2011. *Stata Statistical Software: Release 12*. College Station, TX: StataCorp LP) for analysis. Pairwise comparison for means using paired t-tests were performed to compare the mean difference in monthly and period attendance counts between the two years (2019 considered pre- and 2020 peri-COVID-19 pandemic period). Two-sample test of proportions were computed to compare the hospital RMNCAH services utilisation and maternal and perinatal outcomes before and during the COVID-19 pandemic. P - values ≤0.05 were considered statistically significant. The null hypothesis was that there was no difference in the outcome measures in March to June 2019 compared to March to June 2020. When a statistically significant difference was identified, the null hypothesis was rejected.

### Ethical considerations

No institutional research and ethics review was sought because KHIS data is deidentified open-source and publicly available. Also the use of aggregate data is not human subject research as defined by the World Medical Association’s declaration of Helsinki [22, 23].

## Results

### Overall hospital attendance for RMNCAH services

There were no significant changes in the mean total hospital attendance per month for antenatal care, hospital skilled births, family planning, post-abortion care and immunization services during a 4-month period (March – June 2019) pre-COVID-19 compared with during the equivalent 4-month period peri-COVID-19 pandemic (P>0.05). Even though the changes were not statistically significant, there were reductions in the attendance for antenatal care and immunization services with increments in hospital births, family planning and post-abortion care services (Table 1).

**Table 1:** Monthly hospital attendances during the pre-COVID-19 and peri-COVID-19 periods in Kenya

|  | March - June 2019 |  | March - June 2020 |  |  |
| --- | --- | --- | --- | --- | --- |
| Service | Mean | SD | Mean | SD | P-value |
| Antenatal care | 400,191.3 | 12,700.0 | 384,697.3 | 20,838.6 | 0.251 |
| Hospital births | 98,713.0 | 4,117.0 | 99,634.5 | 3,215.5 | 0.736 |
| Family planning | 431,930.5 | 19,059.9 | 448,168.3 | 31,559.8 | 0.412 |
| Post-abortion care | 3,206.5 | 111.7 | 3,313.0 | 136.7 | 0.273 |
| Pentavalent 1 immunization | 114,701.0 | 3,701.1 | 110,915.8 | 7,209.4 | 0.386 |

It is notable that the trends across the months showed a reduction in hospital attendance in April 2020 for all the hospital services followed by a sustained increase in May and June 2020 for antenatal care, family planning, pentavalent immunization and post-abortion care compared with the similar equivalent pre-COVID-19 period (Fig 2).

**Figure 2:**
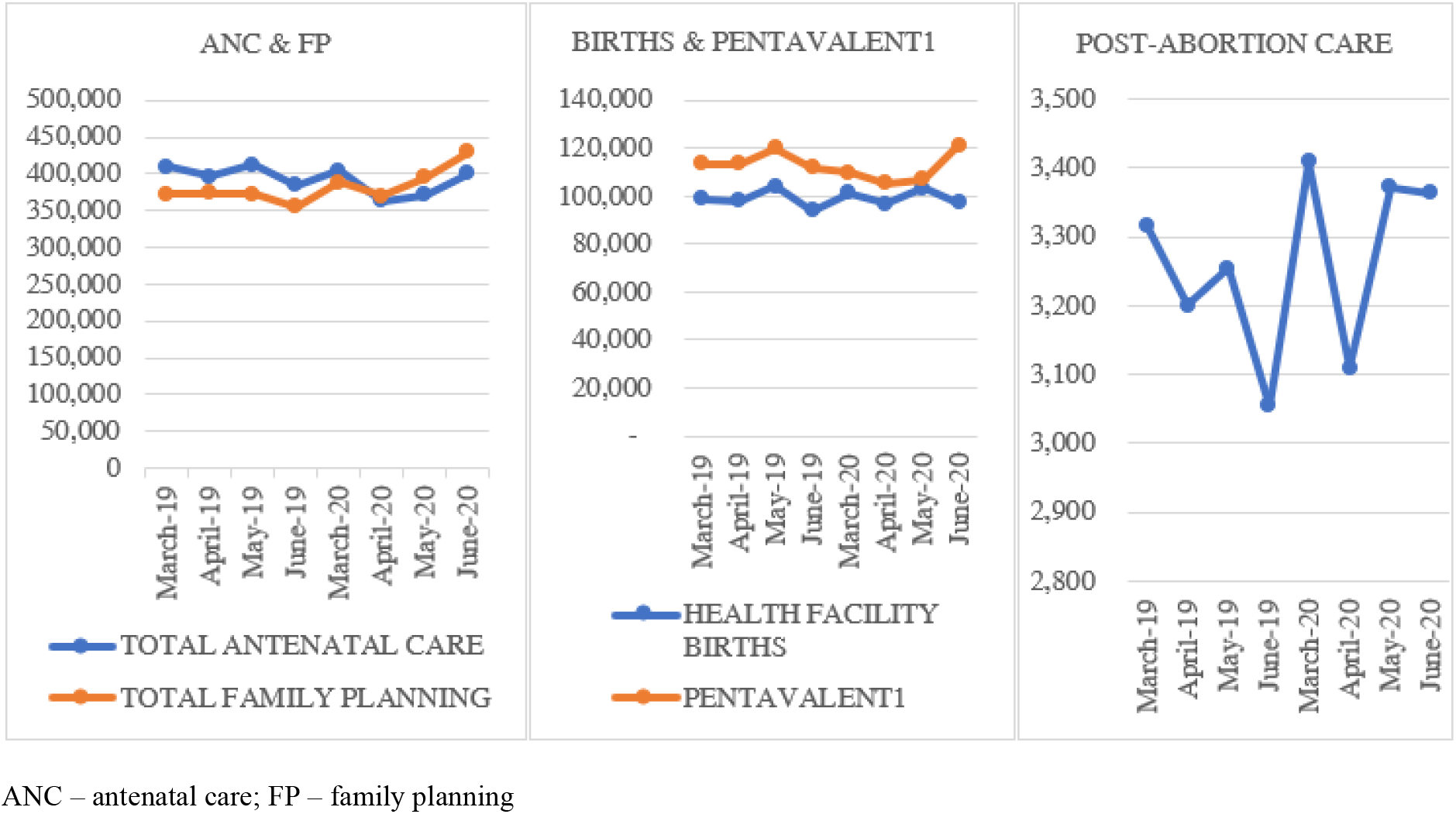
Trends for hospital attendances in equivalent 4-month periods (pre- and peri-COVID-19 pandemic)

### Pregnancy and antenatal care service utilisation

There was a significant reduction in the proportion of adolescents presenting with pregnancy among 10 – 14 years and 15 – 19 years from 0.4% to 0.3% (p< 0.0001) and 8.4% to 7.0% (p< 0.0001) respectively from the pre-COVID-19 to the peri-COVID-19 period. Similar reduction trends were observed for revisiting clients for antenatal care services (69.8% to 67.9%, p< 0.0001) and clients completing four antenatal care visits at the health facilities (18.6% to 17.0%, p< 0.0001). Interestingly, there was a significant increase in the new clients seeking antenatal care services at the health facilities during similar periods (30.2% to 32.1%, p< 0.0001) but also a significant decrease in ANC revisits (69.8% to 67.9%, p< 0.001) although climbing up to March 2019 and March 2020 levels (Table 2 and Figure 1).

**Table 2:** Pre- and peri-COVID-19 pandemic two-sample test of proportions for RMNCAH services utilisation and outcomes in Kenya

| Variable | 2019 |  | 2020 |  | P-value |
| --- | --- | --- | --- | --- | --- |
|  | Total | Proportion | Total | Proportion |  |
| **Pregnancy (n=total ANC clients)** | **(n=1,600,765)** |  | **(n=1,538,789)** |  |  |
| Adolescents 10-14 years with pregnancy | 6,872 | 0.4 | 4,971 | 0.3 | <0.0001 |
| Adolescents 15-19 years with pregnancy | 133,708 | 8.4 | 107,667 | 7.0 | <0.0001 |
| New ANC clients | 482,719 | 30.2 | 493,877 | 32.1 | <0.0001 |
| Re-Visit ANC clients | 1,118,046 | 69.8 | 1,044,912 | 67.9 | <0.0001 |
| Pregnant women completing 4 ANC visits | 298,211 | 18.6 | 261,444 | 17.0 | <0.0001 |
| **FP by client/age (n=total FP clients)** | **n=1,727,722** |  | **n=1,792,673** |  |  |
| FP New clients | 811,464 | 47.0 | 777,119 | 43.3 | <0.0001 |
| FP Revisits | 916,258 | 53.0 | 1,015,544 | 56.6 | <0.0001 |
| Proportion of adolescent FP uptake 10-14 years | 9,017 | 0.5 | 11,592 | 0.6 | <0.0001 |
| Proportion of adolescent FP uptake 15-19 years | 118,667 | 6.9 | 121,581 | 6.8 | 0.0007 |
| Proportion of youth FP uptake (20 – 24 years) | 315,666 | 18.3 | 350,497 | 19.6 | <0.0001 |
| **FP method (n=total women of reproductive age who received a modern contraceptive method)** | **n=1,474,053** |  | **n=1,585,597** |  |  |
| Proportion of E-pills | 25,265 | 1.7 | 25,989 | 1.6 | <0.0001 |
| Proportion of Injectable FP | 858,317 | 58.2 | 988,283 | 62.3 | <0.0001 |
| Proportion of IUCD utilization | 87,461 | 5.9 | 66,794 | 4.2 | <0.0001 |
| Proportion of Implants utilization | 242,706 | 16.5 | 206,299 | 13.0 | <0.0001 |
| Proportion of COC pills | 199,863 | 13.6 | 222,531 | 14.0 | <0.0001 |
| **Post-abortion care services** | **n=12,826** |  | **n=13,252** |  |  |
| Adolescents 10-19yrs accessing PAC services | 2,793 | 21.8 | 2,464 | 18.6 | <0.0001 |
| Women accessing PAC services | 10,033 | 78.2 | 10,788 | 81.4 | <0.0001 |
| **Maternal & Perinatal outcomes (n=total births)** | **n=394,852** |  | **n=398,538** |  |  |
| Caesarean Sections (CS) | 57,541 | 14.6 | 62,815 | 15.8 | <0.0001 |
| Fresh stillbirths (FSB) | 3,634 | 0.9 | 3,883 | 1.0 | 0.0066 |
| Macerated stillbirths (MSB) | 3,805 | 1.0 | 3,959 | 1.0 | 0.0893 |
| Neonatal deaths (NND)<sup>a</sup> | 4,082 | 1.1 | 3,712 | 1.0 | <0.0001 |
| **n=Total maternal deaths** | **n=373** |  | **n=412** |  |  |
| Adolescent (10-19 years) maternal deaths | 23 | 6.2 | 45 | 10.9 | 0.009 |
| Maternal deaths 20+yrs | 350 | 93.8 | 367 | 89.1 | 0.009 |
| Maternal mortality ratio<sup>a</sup> | 96.6<sup>b</sup> |  | 105.8<sup>b</sup> |  | 0.1023 |

a-denominator was total number of live births (385,996 for 2019 and 389,437 for 2020)
b-expressed per 100,000 live births
ANC – antenatal care; FP – family planning; COC – combined oral contraceptives; IUCD – intra-uterine contraceptive device; PAC – post-abortion care

Despite the reduction in proportions of adolescent pregnancy for both the 10 – 14 and 15 – 19 years groups, monthly trends showed a steady rise in numbers of pregnancy in the 15 – 19 years group during the COVID-19 period compared to the pre-COVID period with a flat curve observed in the 10 – 14 years group during similar periods (Table 2 and Table 3).

**Table 3:** Pre – and peri-COVID-19 proportion trends in adolescent RMNCAH service utilisation and outcomes in Kenya

|  | 2019 |  |  |  | 2020 |  |  |  |
| --- | --- | --- | --- | --- | --- | --- | --- | --- |
| Category | MARCH | APR | MAY | JUNE | MARCH | APR | MAY | JUNE |
| Pregnancy (10-14 years) | 0.4 | 0.4 | 0.5 | 0.4 | 0.3 | 0.4 | 0.3 | 0.3 |
| Pregnancy (15-19 years) | 8.4 | 8.3 | 8.3 | 8.4 | 6.7 | 6.8 | 7.2 | 7.3 |
| Pregnancy (10-19 years) | 8.8 | 8.6 | 8.8 | 8.9 | 7.0 | 7.2 | 7.6 | 7.6 |
| Family Planning (10-14 years) | 0.5 | 0.5 | 0.4 | 0.7 | 0.5 | 0.9 | 0.6 | 0.6 |
| Family Planning (15-19 years) | 7.5 | 6.2 | 6.0 | 7.8 | 6.5 | 6.5 | 7.0 | 7.1 |
| Family Planning (10-19 years) | 8.0 | 6.7 | 6.5 | 8.5 | 6.9 | 7.4 | 7.7 | 7.7 |
| Maternal deaths (10-19 years) | 2.3 | 4.8 | 11.5 | 6.0 | 3.6 | 5.9 | 11.9 | 23.0 |

### Hospital births service utilisation

The mean monthly skilled birth attendance rate increased significantly from 68.5% (SD±2.0) to 79.7% (SD±1.2) from the 4-month pre-COVID-19 to peri-COVID-19 period (p< 0.0001). Mean caesarean section births increased significantly from 14,385.3 (SD±689.9) to 15,703.8 (SD±496.8) (p = 0.021) with the overall caesarean section rate increasing significantly from 14.6% to 15.8% (p< 0.0001) during the same periods (Table 2). Comparing the trends for caesarean section rates, there was a steady increase across the peri-COVID-19 period beyond the 10–15% recommended by WHO compared with the pre-COVID rates that were within the WHO recommendations (Fig 3).

**Figure 3:**
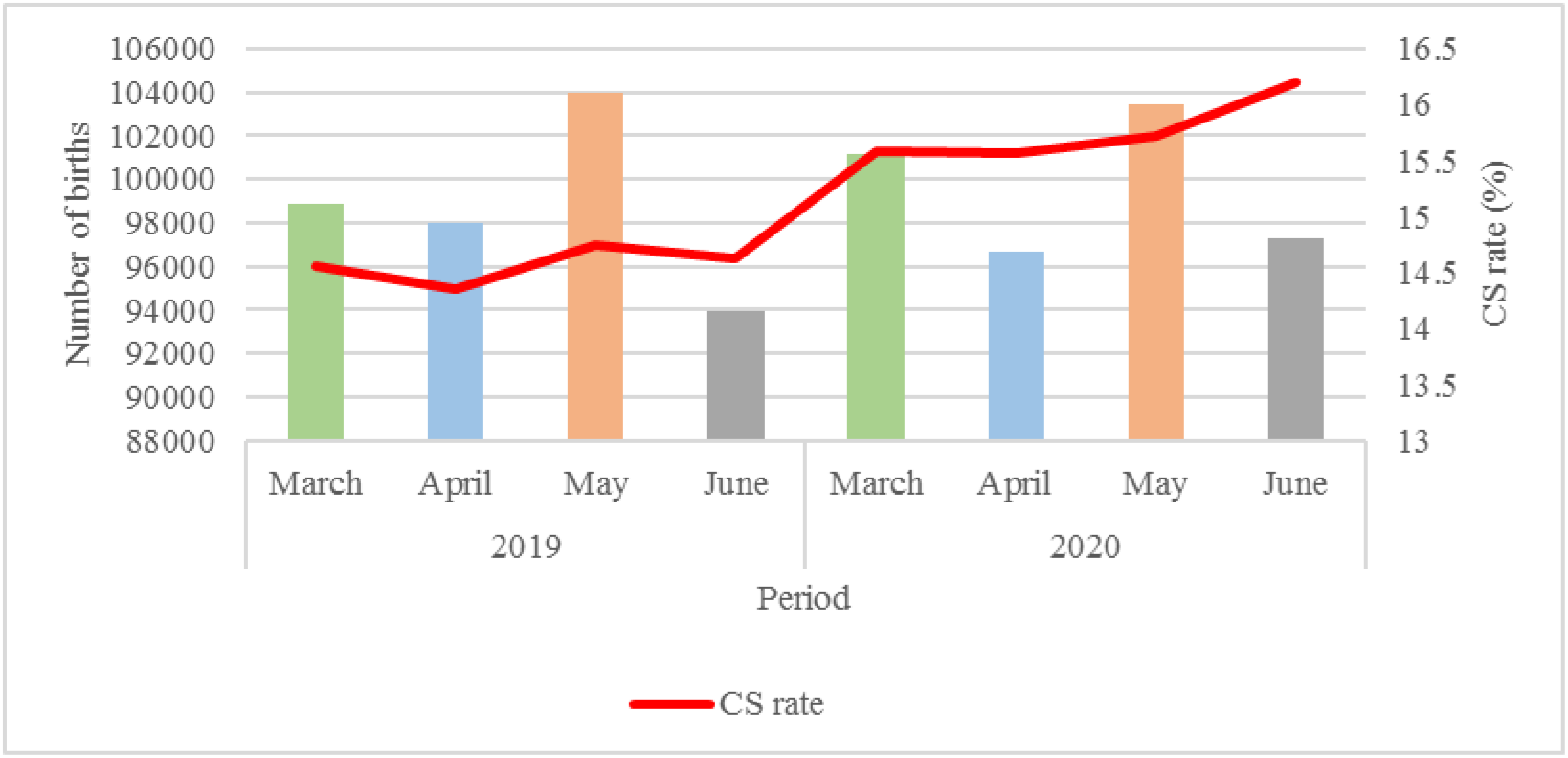
Pre- and peri-COVID-19 trends for total births and CS rates in Kenya

### Family planning and post-abortion care utilisation

There was a significant increase in the proportion of clients revisiting the hospitals for FP services from 53.0% to 56.6% (P< 0.0001) from the pre-COVID-19 to the peri-COVID-19 period. In contrast, a significant reduction was reported in the new clients seeking FP services from 47.0% to 43.3% (P< 0.0001). The proportion of adolescents 10 – 19 years seeking post-abortion care services reduced significantly from 21.8% to 18.6% while the women seeking the same services increased from 72.8% to 81.4% from the pre- to peri-COVID-19 period (p< 0.0001).

Comparing FP services utilisation by age, there was a significant increase in proportion among adolescents 10 – 14 years (0.5% to 0.6%) and youths 20 – 24 years (18.3% to 19.6%) seeking the FP services in the health facilities (p< 0.0001). Comparing by method, injectable FP method was the most utilised both pre- and peri-COVID-19 periods. There were significant increases in the proportion of clients utilising injectable FP methods (58.2% to 62.3%) and combined oral contraceptive pills (13.6% to 14.0%) during similar periods (p< 0.0001). In contrast, there were significant reductions in the proportions of clients utilising emergency contraceptive pills (1.7% to 1.6%), intrauterine devices (5.9% to 4.2%) and implants (16.5% to 13.0%) during the same periods (p< 0.0001) (Table 2). Comparing trends over different data points, there was a steady rise in adolescents 15 – 19 years and overall 10 – 19 years utilising FP services across all the 4-months peri-COVID-19 period (Table 3).

### Maternal and perinatal mortality

During the 4-months periods, the proportion of adolescent (10 – 19 years) maternal deaths significantly increased from 6.2% to 10.9% from the pre- to peri-COVID-19 period (P< 0.009). During the same period, there was a significant increase in the proportion of fresh stillbirths from 0.9% to 1.0% (P = 0.0066). However, there was a significant reduction in the proportion of neonatal deaths from 1.1% to 1.0% (P<0.0001) with no changes in the proportions for macerated stillbirths during the two periods (Table 2). Like for trends across the months for the other indicators, there was a steady rise in fresh stillbirths, macerated stillbirths and adolescent maternal deaths during the four peri-COVID-19 months.

## DISCUSSION

This study has demonstrated the overall impact of COVID-19 pandemic on utilisation and maternal and perinatal outcomes during the first four months, when strict measures were in place to limit community spread in a low – and middle-income setting.

Overall, there was no significant change in the mean total hospital attendance for all the RMNCAH services – antenatal care, skilled birth, family planning, post-abortion care and immunization services during the 4-month peri-COVID-19 period as per the earlier global predictions [18, 19]. However, the trends across the services showed a dip in April 2020 before eventually rising in May and June reaching and/or even surpassing the 4-month pre-COVID-19 period. This could be attributed to a well-coordinated and centralised risk communication strategy by the government first by decreeing that health services were critical essential services to be accessed by all. Secondly, the centralised communication campaigns through daily ministerial briefings (through radio and television) and other social media platforms helped to dispel any fears from the public in seeking hospital care for the critical essential services. Experts including the WHO, London School of Hygiene and Tropical Medicine and UNICEF recommend that risk communication is a critical component of the response to COVID-19 as is with other epidemics [24, 25]. Such communication helps the public make the right decisions about how to protect themselves, when to seek care, and to avoid contributing to panic about the disease and its effects. This approach manages the ‘infodemic’ (an excess of information that makes it hard to know what’s trustworthy and what’s not) and maintains trust in public health authorities that is critical to any ongoing management of the outbreak [24].

There was a general reduction in new ANC clients, the uptake of long-term family planning methods and an increase in the short-term contraceptive utilization during the peri-COVID-19 period. It appears like current FP users were more motivated to continue using compared with never-users in the initial months of the pandemic. Evidence suggests that first discussion with a health care provider is a key determinant for uptake of long acting and permanent contraceptive methods among women of reproductive age [26–28]. The injectable contraception was the most prevalent FP method used during the peri-COVID-19 period. Besides, this method had the highest increase in uptake during the period. Injectable and other short acting family planning methods remain the most dominant among women of reproductive age in sub-Saharan Africa [29, 30]. Women prefer this method as it is considered private: no one else can tell that a woman is using contraception [31]. Evidence from 33 sub-Saharan countries showed that young women (15–24 years) prefer short-term methods obtained from limited-capacity, private providers, compared with older women. Besides, the use of long-term methods among young women is low, but among those users, more than 85 percent receive the service from a public sector source [32].

A decrease in the unmet need for FP and increase in utilisation of PAC service in the first 4 months of the pandemic compared to the same period a year earlier is encouraging. Maintaining adequate stock levels and supply chain throughout the year, is essential to maintaining this trend. Although, globally, there are anticipated shortages of contraception, we have not analysed the FP supply chain in Kenya during this period [33]. It is unclear what the impact of the pandemic was on the supply chain for various FP commodities in Kenya, this may also have contributed to the low uptake of implants in the first 4 months of the pandemic in Kenya.

An interesting situation was seen with adolescents accessing RH/MNH services: there were more adolescent pregnancies, increased uptake of FP services, reduced uptake of PAC services but increased adolescent maternal deaths. Closure of schools could have a part to play in this trend as majority of this age group is school going and therefore kept engaged with the rigorous education routines that have been suspended. A research conducted by Plan International in nine counties of Kenya in 2019 showed that 98 percent of pregnant girls were not in school, and 59 percent of the pregnancies among girls aged 15–19 years were unintended. Furthermore, 45 percent of severe abortion complications were also reported among adolescent girls [34]. Evidence shows that pregnancy before the age of 18 years increases the health risks for the mother and her baby [35]. Clearly the effect of increased FP uptake amongst adolescents in terms of health outcomes will not be visible in the short term but further investigation of adolescent maternal deaths recorded during this period is required to understand the underlying causes of deaths, associated and contributing factors related to these deaths. This is more so because the there was a significant reduction of maternal deaths in women aged 20 and above compared to adolescents in the same period. This is critical to ensuring that the predicted additional maternal deaths are avoided during this pandemic and during similar outbreaks in future.

The caesarean section rates in the peri-COVID-19 period are steadily rising beyond the WHO recommendation of not more than 10–15 percent as rates higher than 10 percent are not associated with reductions in maternal and newborn mortality rates [36]. The rising caesarean section rates are consistent with evidence from a systematic review among women who had COVID-19 [37]. According to WHO, Royal College of Obstetricians and Gynaecologists (RCOG) and Kenya MOH COVID-19 guidelines, COVID-19 infection is not an indication for caesarean section and therefore the rising rate of caesarean section needs to be monitored and investigated [25, 38, 39]. It would be expected that this rise should translate to a reduction in the fresh stillbirths in the facilities. However, there is an unexpected rise in the fresh stillbirths which could reflect falling standards of quality of care during labor and childbirths in the health facilities during the pandemic. Dawn-to-dusk curfews, related fear to police enforcers of the curfews (leading to interrupted transport and support systems) and lockdowns presented a worrying period for pregnant women disrupting access to antenatal and skilled birth services [40]. This could easily lead to delays in accessing skilled health services further complicating pregnancies/labor which could have contributed to the rise in stillbirths in the country. Consequently, evidence from Kenya, Uganda and Tanzania showed that worries among midwives in provision of essential health care services due to poor infrastructure, lack of/inadequate personal protective equipment and fear of contracting the disease could have contributed to the worsened trend for stillbirths in the country [40–42]. The exact nature of the delays experience by women when they reach health facilities needs further investigation, this could potentially include but not limited to delays in providing care due to reduced number of health workers, poor preparedness of facilities to operate during the pandemic, and increased frequency of disrespectful maternity care [43, 44].

For any reduction in preventable maternal and perinatal mortalities, WHO emphasizes that pregnant women must be able to access the right care at the right time [45]. Therefore, resources (human capacity) and a supporting environment (drugs and supplies and a functional referral system) need to be guaranteed to ensure access to skilled health personnel for deliveries and emergency obstetric care in the face of increased COVID-19 threats. The rising macerated stillbirths indicate that women are likely not seeking hospital care early enough and/or they are seeking care from unskilled health personnel in the communities [34, 40]. Public health responses to COVID-19 should leverage intersectional, human rights centred frameworks and community-driven approaches to sufficiently prevent complex health adversities for women, adolescent girls, and vulnerable populations [46].

Our study has strengths and limitations. The use of a comparison period enabled testing the hypothesis on whether the COVID-19 pandemic had any impact on the utilization of RMNCAH services and maternal and perinatal outcomes. However, the use of DHIS2 data poses key data quality challenges including inaccurate and incomplete reporting that are prevalent in low and middle-income countries [47–50]. The short period reviewed during the peri-COVID-19 pandemic represents the period when the burden of the pandemic was not at the peak in the country compared to the transmission trends as experienced in other European and American countries at similar periods and therefore the results should be interpreted in context. Further evaluations at 6 and 12 months may be required to validate the outcomes.

Based on the earlier models, the predictions in reduction of access and utilization of essential RMNCAH services and worsening maternal and perinatal mortality did not hold at four months. The concerted efforts by the government in managing the communication and messaging during the pandemic played a key difference although further evaluations may be required to clearly assess the impact of the pandemic on RMNCAH services and maternal and perinatal outcomes.

## Conclusion

Findings demonstrate that the earlier gains achieved in family planning, pregnancy and skilled child births and adolescent sexual and reproductive health are likely to be watered down by the COVID-19 pandemic. The rising caesarean sections with no improvement in maternal and perinatal outcomes require further monitoring and investigation. While the overall picture for maternal mortality ratio is reassuring, a significant increase in maternal deaths amongst adolescents is of concern. If unchecked, there is a likelihood of worsening of adolescent maternal deaths and stillbirths. Further research urgently needed to understand the reasons for increased adolescent maternal mortality, reduced uptake of long-term FP methods, and increased still birth rate. This will be critical to developing policies and interventions to prevent additional maternal deaths during this pandemic and in future infectious disease outbreaks.

## What is already known on this topic

COVID-19 like other pandemics, is projected to result in low access and utilization of reproductive, maternal and child health services
Women and children are the most affected during the crises caused by pandemics

## What this study adds

The COVID-19 has contributed to a high utilization of short-term family planning methods (injectables and pills) with a reduction in long-term methods (implants and intra-uterine devices).
There are rising cases of caesarean section rates beyond the WHO recommended 10–15% with no improvement in maternal and perinatal mortality rates.
There is rising teenage pregnancy rates with a rising adolescent maternal mortality rates but decreased maternal mortality amongst women aged 20 years and above.

## Data Availability

Data is available from the corresponding author upon reasonable request.

## Acknowledgement

This paper has been developed under the Reducing Maternal and Newborn Deaths programme as part of efforts to contribute to understanding the impact of the COVID-19 on maternal and newborn health. The authors acknowledge the United Kingdom Department for International Development (DfID) through the Reducing Maternal and Newborn Deaths programme supporting the reduction of maternal and perinatal deaths in Kenya focusing at the national and subnational (counties) level.

## Authors’ contributions

DS and CA conceptualized the idea and designed the study. DS performed data extraction, cleaning, analysis and interpretation of the results, drafted the primary manuscript, reviewed and prepared it for publication. CA interpreted the results, reviewed and prepared the manuscript for publication. IN, SG, OM, ME, PG and LN participated in the design of the study procedures and reviewed the final manuscript. All the authors have read and approved the final manuscript.

## References

1. World Health Organization. WHO Director-General’s opening remarks at the media briefing on COVID-19 - 11 March 2020. Geneva: World Health Organization. Available from: https://www.who.int/dg/speeches/detail/who-director-general-s-opening-remarks-at-the-media-briefing-on-covid-19---11-march-2020.

2. World Health Organization. Coronavirus disease (COVID-19) Situation Report – 162 World Health Organization; 2020. Available from: https://www.who.int/docs/default-source/coronaviruse/20200630-covid-19-sitrep-162.pdf?sfvrsn=e00a5466_2.

3. Euronews. Coronavirus second wave? Which countries in Europe are experiencing a resurgence of cases? 2020. Available from: https://www.euronews.com/2020/08/27/is-europe-having-a-covid-19-second-wave-country-by-country-breakdown.

4. DW. Coronavirus pandemic: Is the second wave already here? 2020. Available from: https://www.dw.com/en/coronavirus-second-wave/a-54429614.

5. New Scientist. Coronavirus: Second wave hits Asia as global cases continue to soar 2020. Available from: https://www.newscientist.com/article/2249982-coronavirus-second-wave-hits-asia-as-global-cases-continue-to-soar/.

6. World Health Organization. Maintaining essential health services: operational guidance for the COVID-19 context: Interim guidance 1 June 2020: World Health Organization; 2020. Available from: https://www.who.int/publications/i/item/covid-19-operational-guidance-for-maintaining-essential-health-services-during-an-outbreak.

7. Vindegaard N, Benros ME. COVID-19 pandemic and mental health consequences: systematic review of the current evidence. Brain, Behavior, and Immunity. 2020.

8. United Nations Educational SaCOU. Taking action to end domestic violence during pandemics 2020. Available from: https://en.unesco.org/news/taking-action-end-domestic-violence-during-pandemics.

9. World Health Organization. Managing epidemics: Key facts about major deadly diseases. Geneva: World Health Organization 2018. Available from: https://www.who.int/emergencies/diseases/managing-epidemics/en/.

10. World Health Organization. Strategy and planning: strategic preparedness and response plan. Geneva: World Health Organization; 2020. Available from: https://www.who.int/emergencies/diseases/novel-coronavirus-2019/strategies-and-plans.

11. World Health Organization. COVID-19: Operational guidance for maintaining essential health services during an outbreak: interim guidance, 25 March 2020. Geneva: World Health Organization; 2020. Available from: https://apps.who.int/iris/handle/10665/331561.

12. World Health Organization. Community-based health care, including outreach and campaigns, in the context of the COVID-19 pandemic: interim guidance, May 2020. Geneva: World Health Organization, United Nations Children’s Fund; 2020. Available from: https://apps.who.int/iris/handle/10665/331975.

13. Elston J, Cartwright C, Ndumbi P, Wright J. The health impact of the 2014–15 Ebola outbreak. Public health. 2017;143:60–70.

14. Elston J, Moosa A, Moses F, Walker G, Dotta N, Waldman RJ, et al. Impact of the Ebola outbreak on health systems and population health in Sierra Leone. Journal of Public Health. 2016;38(4):673–8.

15. Sochas L, Channon AA, Nam S. Counting indirect crisis-related deaths in the context of a low-resilience health system: the case of maternal and neonatal health during the Ebola epidemic in Sierra Leone. Health policy and planning. 2017;32(suppl_3):iii32-iii9.

16. Bietsch K, Williamson J, Reeves M. Family planning during and after the West African Ebola crisis. Studies in Family Planning. 2020;51(1):71–86.

17. Camara BS, Delamou A, Diro E, Béavogui AH, El Ayadi AM, Sidibé S, et al. Effect of the 2014/2015 Ebola outbreak on reproductive health services in a rural district of Guinea: an ecological study. Transactions of The Royal Society of Tropical Medicine and Hygiene. 2017;111(1):22–9.

18. Roberton T, Carter ED, Chou VB, Stegmuller AR, Jackson BD, Tam Y, et al. Early estimates of the indirect effects of the COVID-19 pandemic on maternal and child mortality in low-income and middle-income countries: a modelling study. The Lancet Global Health. 2020.

19. Riley T, Sully E, Ahmed Z, Biddlecom A. Estimates of the potential impact of the COVID-19 pandemic on sexual and reproductive health in low-and middle-income countries. Int Perspect Sex Reprod Health. 2020;46:46.

20. Polit DF, Beck CT. Nursing research: Principles and methods: Lippincott Williams & Wilkins; 2004.

21. Ministry of Health (Kenya). COVID-19 OUTBREAK IN KENYA: DAILY SITUATION REPORT – 105: Ministry of Health (Kenya); 2020. Available from: https://www.health.go.ke/wp-content/uploads/2020/07/Kenya-COVID-19-SITREP-0105-30-Jun-2020.pdf.

22. World Medical Association. World medical association declaration of helsinki: Ethical principles for medical research involving human subjects. JAMA. 2013;310(20):2191–4.

23. Kenya Health Information System [Internet]. 2020 [cited 3rd Aug 2020]. Available from: https://hiskenya.org/dhis-web-pivot/.

24. London School of Hygiene & Tropical Medicine. COVID-19: Tackling the Novel Coronavirus 2020. Available from: https://www.futurelearn.com/courses/covid19-novel-coronavirus/1.

25. World Health Organization. Maintaining essential health services: operational guidance for the COVID-19 context: Interim guidance 1st June 2020. Geneva: World Health Organization; 2020. Available from: https://apps.who.int/iris/handle/10665/332240.

26. Melka AS, Tekelab T, Wirtu D. Determinants of long acting and permanent contraceptive methods utilization among married women of reproductive age groups in western Ethiopia: a cross-sectional study. Pan African Medical Journal. 2015;22(1).

27. 27. Teferra AS, Wondifraw AA. Determinants of long acting contraceptive use among reproductive age women in Ethiopia: evidence from EDHS 2011. 2015.

28. Habtamu A, Tesfa M, Kassahun M, Animen S. Determinants of long-acting contraceptive utilization among married women of reproductive age in Aneded district, eterminants of long-acting contraceptive utilization among married women of reproductive age in Aneded district, Ethiopia: a case-control study. BMC Research Notes.2019;12(1):433.

29. United Nations, Department of Economic and Social Affairs, Population Division. Contraceptive Use by Method 2019: Data Booklet 2019. Available from: https://www.un.org/development/desa/pd/sites/www.un.org.development.desa.pd/files/files/documents/2020/Jan/un_2019_contraceptiveusebymethod_databooklet.pdf.

30. Ross JA, Agwanda AT. Increased use of injectable contraception in sub-Saharan Africa. African journal of reproductive health. 2012;16(4):68–80.

31. World Health Organization Department of Reproductive Health and Research (WHO/RHR) and Johns Hopkins Bloomberg School of Public Health/Center for Communication Programs (CCP) KfHP. Family Planning: A Global Handbook for Providers (2018 update). Baltimore and Geneva: WHO and CCP; 2018. Available from: https://www.fphandbook.org/sites/default/files/global-handbook-2018-full-web.pdf.

32. Radovich E, Dennis ML, Wong KL, Ali M, Lynch CA, Cleland J, et al. Who meets the contraceptive needs of young women in sub-Saharan Africa? Journal of Adolescent Health. 2018;62(3):273–80.

33. Chris P. Opinion: how will COVID-19 affect global access to contraceptives—and what can we do about it? Devex. 2020.

34. Plan International. COVID-19: LOCKDOWN LINKED TO HIGH NUMBER OF UNINTENDED TEEN PREGNANCIES IN KENYA 2020. Available from: https://plan-international.org/news/2020-06-25-covid-19-lockdown-linked-high-number-unintended-teen-pregnancies-kenya.

35. Unicef W, Unesco, Unfpa, Undp, Unaids, WFP and the World Bank,. Facts for life: United Nations Children’s Fund, New York; 2010. Available from: http://www.factsforlifeglobal.org/resources/factsforlife-en-full.pdf.

36. World Health Organization. WHO Statement on Caesarean Section Rates. Geneva: World Health Organization; 2015. Available from: https://apps.who.int/iris/bitstream/handle/10665/161442/WHO_RHR_15.02_eng.pdf?sequence=1.

37. Smith V, Seo D, Warty R, Payne O, Salih M, Chin KL, et al. Maternal and neonatal outcomes associated with COVID-19 infection: A systematic review. Plos one. 2020;15(6):e0234187.

38. Royal College of Obstetricians and Gynaecologists (RCOG). Coronavirus (COVID-19) Infection in Pregnancy: Information for healthcare professionals. London: Royal College of Obstetricians and Gynaecologists (RCOG); 2020. Available from: https://www.rcog.org.uk/globalassets/documents/guidelines/2020-07-24-coronavirus-covid-19-infection-in-pregnancy.pdf.

39. Ministry of Health (Kenya). KENYA COVID19 RMNH GUIDELINES: A Kenya Practical Guide for Continuity of Reproductive, Maternal, Newborn and Family Planning Care and Services in the Background of COVID-19 Pandemic MOH, April 2020. Nairobi: Ministry of Health (Kenya); 2020. Available from: https://www.health.go.ke/wp-content/uploads/2020/04/KENYA-COVID19-RMNH.pdf.pdf.pdf.

40. Alliance TWR. COVID-19 Curfew Restrictions Impact Reproductive, Maternal, and Newborn Health and Rights Worldwide. News [Internet]. 2020 20th August 2020. Available from: https://www.whiteribbonalliance.org/2020/06/08/covid-19-curfew-restrictions-impact-women-andnewborns-worldwide/.

41. AP News. Pregnant women at risk of death in Kenya’s COVID-19 curfew20th August 2020. Available from: https://apnews.com/2e1a7d8b8401e4c06df52085994cf4ba.

42. Pallangyo E, Nakate MG, Maina R, Fleming V. The impact of covid-19 on midwives’ practice in Kenya, Uganda and Tanzania: A reflective account. Midwifery.2020.

43. Delamou A, Hammonds RM, Caluwaerts S, Utz B, Delvaux T. Ebola in Africa: beyond epidemics, reproductive health in crisis. Lancet. 2014;384(2105):62364–3.

44. Wilhelm JA, Helleringer S. Utilization of non-Ebola health care services during Ebola outbreaks: a systematic review and meta-analysis. Journal of global health. 2019;9(1).

45. World Health Organization. Pregnant women must be able to access the right care at the right time, says WHO. WHO Newsroom [Internet]. 20th August 2020. Available from: https://www.who.int/en/news-room/detail/07-11-2016-pregnant-women-must-be-able-to-access-the-right-care-at-the-right-time-says-who.

46. Hall KS, Samari G, Garbers S, Casey SE, Diallo DD, Orcutt M, et al. Centring sexual and reproductive health and justice in the global COVID-19 response. The Lancet. 2020;395(10231):1175–7.

47. Rowe AK, Kachur SP, Yoon SS, Lynch M, Slutsker L, Steketee RW. Caution is required when using health facility-based data to evaluate the health impact of malaria control efforts in Africa. Malaria journal. 2009;8(1):209.

48. Bhattacharya AA, Umar N, Audu A, Felix H, Allen E, Schellenberg JR, et al. Quality of routine facility data for monitoring priority maternal and newborn indicators in DHIS2: A case study from Gombe State, Nigeria. PLoS One. 2019;14(1):e0211265.

49. Hagel C, Paton C, Mbevi G, English M. Data for tracking SDGs: challenges in capturing neonatal data from hospitals in Kenya. BMJ Global Health. 2020;5(3):e002108.

50. Nielsen P. Advancing health information systems: Experiences from implementing DHIS 2 in Africa. 2013.

